# The Change of Screen Time and Screen Addiction, and their Association with Psychological Well-being During the COVID-19 Pandemic: An Analysis of US Country-Wide School-Age Children and Adolescents Between 2018 and 2020

**DOI:** 10.1101/2023.03.20.23287490

**Authors:** Helena T. Wu, Jiandong Li, Amy Tsurumi

## Abstract

Previous studies on screen use and children’s mental health during the Coronavirus Disease 2019 (COVID-19) pandemic either focused only on the timeframe during the pandemic, or only on children previously reporting COVID-related severe family economic hardship or worries. Instead, we used a large sample (n=63,211) of the National Survey of Children’s Health (NSCH) years 2018-20 to analyze changes in the trends of recreational screen device use before, versus during the COVID-19 pandemic, and associations with psychological well-being, widely among school-age children (6-17 year-olds) across the US. We assessed recreational screen use, instead of overall use including both instructional and recreational use, and developed psychological well-being issue scores to evaluate the associations among the pandemic, recreational screen use, and psychological well-being states. We found an increase in the prevalence of screen overuse/addiction and psychological well-being issues during the pandemic compared to the years prior, detected an association between the pandemic and psychological well-being issue scores (p <0.01 across all models), and observed increased magnitude of association between recreational screen overuse/addiction and mental health during the pandemic year (p <0.01 across all models). Further studies on elucidating and addressing the specific aspects of the pandemic that contribute to these associations are critical.

## Introduction

Previous studies have shown that children and adolescents’ psychological well-being (hereinafter referred to as children and children’s well-being) is negatively associated with screen time.^1,2,3,4,5,6^ The recreational use of video and online games, television, internet, and smartphones, in addition to the increasing educational-based usage of online platforms, has resulted in an increase in overall screen time usage, especially during the COVID-19 pandemic year of 2020.^7,8^ As the pandemic forced most schools to cease face-to-face instruction and instead implement distance learning, school-age children were generally exposed to screens for longer amounts of time. Previous studies have evaluated the impact of the COVID-19 pandemic on screen time and its associations with mental health and well-being conditions.^9,10^ However, these studies evaluated associations during the pandemic only, and a study comparing the relation between screen time and well-being in the United States (US) over time, before and during the pandemic is lacking. Moreover, these studies focused on those having reported severe family and economic impact of COVID-19^9^ or “COVID-related worry”^10^, and within narrow age ranges, and thus a study more widely across all school-age children would be informative. Thus, we investigated trends before and during the pandemic in order to assess whether the pandemic itself is a major event that impacted the state of well-being of school-age children widely across the US overall, in relation to their screen time usage.

The COVID-19 pandemic provides a unique opportunity to investigate a time when children, in general, reached the greatest screen time usage hours across the entire US. Among various extreme screen usages, the American Psychiatric Association (APA) classifies internet gaming as an impulse disorder^11^. Moreover, in 2018, the World Health Organization included gaming disorder in the 11th revision of the International Classification of Diseases^12^. On the other hand, screen addiction from the use of smartphones and other electronic devices has not yet been classified as a clinical disorder, even though the harms of screen overuse specifically related to these devices are well documented.^13,14^ One difficulty in defining addiction disorder from smartphones and other electronic devices, is that their use can result in both positive and negative impact.^15^ Although there is currently no established criteria for defining screen addiction specifically for those devices, excessive daily recreational use, similar to the extent of overuse among those with gaming disorders, may be consistent with symptoms of addiction. The pandemic invariably increased hours spent on the screen due to school and school-related work having been conducted exclusively online. However, significant recreational screen use outside of school-related online instruction during the pandemic could be especially alarming, as it would indicate that a child is constantly on the screen throughout the day, and such excessive screen time could be consistent with screen addiction. Thus, in our study, we specifically assessed recreational (*i.e*. not school-related) screen time. Since an established definition of screen addiction does not exist, in this study, we evaluated the effect of reported excessive recreational screen overuse, and considered it interchangeably with addiction. The objective of this study was to explore the impact of the pandemic on recreational screen overuse/addiction, and its association with various measures of children’s psychological well-being, using a large sample of school-age children widely across the US. Moreover, we explored whether the trends and association have evolved during the year before versus during the pandemic, and the impact of the pandemic itself on strengthening the link between screen time and addiction on psychological well-being.

## Methods

### Study subjects

In 2016, the Census Bureau initiated the National Survey of Children’s Health (NSCH) data collection by administering mail and web-based surveys, instead of the earlier method by telephone.^16^ It annually sends out the questionnaire to randomly chosen children aged 0-17 years, or their caregiver, in 50 states and the District of Columbia, and each household has one randomly chosen respondent. The original dataset is made publicly available, and includes 50,212 observations for 2016, 21,599 for 2017, 30,530 for 2018, 29,433 for 2019, and 42,777 for 2020.

Since the objective of our study is to investigate school-age children, we included only children aged 6-17 years. For study years 2016 and 2017, the screen time definition is different from that of 2018 and beyond. In 2016 and 2017, the NSCH collected computer and TV hours separately, and thus, the total screen time can be ascertained by adding the computer hours and TV hours. The average screen time for 2016 and 2017 was found to be 3.05 hours, with a range between 0 and above 8 hours. Starting in 2018, the NSCH combined the survey question on computer and TV hours, and collected screen time as one measure. The mean screen time between 2018 and 2020 was found to be 2.18 hours with a range between 0 and above 4 hours (these averages were calculated using the original dataset before data filtering), showing a notable difference from the 2016 and 2017 study. Therefore, to maintain consistency, we included data from years 2018, 2019, and 2020 only.

We further removed observations of respondents with reported autism, blindness, cerebral palsy, deafness, Down syndrome, developmental delay, epilepsy, or intellectual disabilities, as psychological well-being measures and screen time behavior may require different considerations among this group. Furthermore, we removed observations of respondents with missing screentime or psychological well-being symptoms observations. In summary, the total number of respondents in this study is 63,211, including 18,746 for 2018, 18,212 for 2019, and 26,253 for 2020 (Supplementary Figure S1).

### Variables and measures used

#### Description of the NSCH data obtained

Details on the NSCH survey variable names according to the code book, questions used to collect data, and descriptions of our study’s recoded data used for subsequent analyses are provided in Supplementary Table S1. We also obtained the 3-year weight for 2018-202 from the NSCH’s Guide to Multi-Year Analysis, and the adjusted weight was used in all the statistical analyses.

#### Subject demographics

We obtained the data on age, race, ethnicity, sex, and poverty ratio. We computed the poverty ratio used in the analyses by averaging across the six imputed poverty ratio values in accordance with the NSCH’s Guide to Analysis with Multiply Imputed Data.

#### Recreational screen time measure

Reported hours that a child reported having spent on screens recreationally, including TV, computer, cellphone, or other electronic devices to watch programs, play games or use social media on most weekdays was used (NSCH data variable, SCREENTIME). The reported value ranges between 0.5 and 4, where 0.5 represents the average screen time of less than 1 hour, and 4 indicates four or more hours. As previously reported^1^, the relationship between children’s psychological well-being and screen time is nonlinear, and low screen usage (<1 hour) appears to be beneficial for children’s psychological well-being (Supplementary Figure S2). Thus, in order to specifically characterize the adverse effect of screen time on psychological well-being, for the regression analysis, we only included observations with at least one hour of screen usage every weekday, leaving a total of 50,527 observations.

#### Screen addiction measures

We considered reported recreational screen time during a weekday of 4 or more hours to be an indicator of screen addiction (variable *Addiction*, yes or no). Considering school hours (8 hours), sleep hours (8 hours), and commuting, eating and personal time (3-4 hours) during a typical school day, there are approximately 4-5 hours remaining for the day. If a child spends all the available 4-5 hours on the screen, there is a reasonable concern about screen overuse which may be indicative of addiction. This is especially the case during the COVID-19 pandemic, where, in addition to spending all school-related hours on the screen, a child spending 4 hours or more hours on recreational screen a day suggests that they are using the screen if they are not eating or sleeping.

#### Psychological well-being variables

In a previous study, psychological well-being was described as a broad concept to reflect various factors including “emotional stability, positive interpersonal relationships, self-control, and indicators of flourishing as well as diagnoses of mood disorders such as anxiety or depression.”^1^ Accordingly, we considered the following NSCH study variables as well-being factors: ability to remain calm and in control when challenged (K7Q85_R), argues too much (K7Q70_R), has difficulty making or keeping friends (MAKEFRIEND), works to finish tasks they have started (K7Q84_R), shows interest and curiosity (K6Q71_R), is difficult to care for (K8Q31), has ever been diagnosed by healthcare professionals with depression (K2Q32A) or anxiety (K2Q33A). The original study variables were recoded as dichotomous variables that indicate issues with well-being for developing psychological well-being proxy measures and downstream analyses (Supplementary Table S1).

#### Psychological well-being proxy measures

A child with lower psychological well-being is expected to have one or more of the above related psychological symptoms described in Supplementary Table S1. In order to describe well-being conditions, we constructed two psychological well-being issue (WBI) scores as described below, and summarized in Supplementary Table S2:

##### WBI1 (well-being issue score 1)

WBI1 was calculated by adding six of the psychological well-being symptom dichotomous variables (*Not Calm, Argues Too Much, Difficult to Make Friends, Does Not Finish Tasks, Not Curious*, and *Difficult to Care*), which are expected to be related to depression and anxiety. WBI1 is a composite score ranging from 0 to 6, where the higher the value is, the lower the psychological well-being.

##### WBI2 (well-being issue score 2)

WBI2 is a dichotomous variable with a value of 1 if any one of the eight well-being symptom category variables is 1 (*Not Calm, Argues Too Much, Difficult to Make Friends, Does Not Finish Tasks, Not Curious, Difficult to Care, Depression*, or *Anxiety*), otherwise is 0. These eight variables are expected to be distinct from one another and the WBI2 score is expected to reflect well-being status generally.

#### Multivariate regression models

Various multivariate Generalized Least Square (GLS) models for WBI1 (Model 1 and Model 2), or logistic regression models for WBI2 (Model 3 and Model 4), were constructed as described below. For each of the four models, either screen time in hours (*Screen Time*) or screen addiction, defined as recreational screen time ≥ 4 hours a day (*Addiction*) was included in the model as the main effect to be assessed. Compared to Model 1 and Model 3, Model 2 and Model 4 include two additional variables: a dummy variable to represent the pandemic year 2020 (*Pandemic 2020*), and an interaction term between the pandemic year 2020 variable and screen time or addiction (*Screen Time/Addiction*Pandemic 2020*), to capture the effect of recreational screen time on well-being issue outcomes depending on whether or not it was the pandemic year. As conventionally conducted in a multi-level model, the centralized and standardized screen time was used to compute the interaction term. The age variable ranged from 6 to 17 years old, and the age categorical variable was defined as elementary school-age children (6-10 years old), middle school-age children (11-13 years old), and high school-age children (14-17 years old) used as the reference group. Categorical variables for sex (female as reference), race (White as reference), and ethnicity (not Hispanic or Latino as reference) were used.

### GLS models for the WBI1 score outcome

#### Model 1

WBI1_Model1_ = *α* + *β*_*1*_ Screen Time/Addiction + *β*_*2*_ Age + *β*_*3*_ Sex-male + *β*_*4*_ Race-Black

+ *β*_*5*_ Race-American Indian or Alaska native + *β*_*6*_ Race-Asian

+ *β*_*7*_ Race-Hawaiian or Pacific Islander + *β*_*8*_ Race-Other + *β*_*9*_ Race-Two or More Races

+ *β*_*10*_ Ethnicity-Hispanic or Latino + *β*_*11*_ Poverty Ratio + *ε*

#### Model 2

WBI1_Model2_ = *α* + *β*_*1*_ Screen Time/Addiction + *β*_*2*_ Pandemic 2020 + *β*_*3*_ Screen Time/Addiction*Pandemic 2020

+ *β*_*4*_ Age + *β*_*5*_ Sex-male + *β*_*6*_ Race-Black + *β*_*7*_ Race-American Indian or Alaska native

+ *β*_*8*_ Race-Asian + *β*_*9*_ Race-Hawaiian or Pacific Islander + *β*_*10*_ Race-Other

+ *β*_*11*_ Race-Two or More Races + *β*_*12*_ Ethnicity-Hispanic or Latino + *β*_*13*_ Poverty Ratio + *ε*

### Logistic regression models for the WBI2 score outcome

#### Model 3

WBI2_Model3_ = *α* + *β*_*1*_ Screen Time/Addiction + *β*_*2*_ Elementary School + *β*_*3*_ Junior School + *β*_*4*_ Sex-male

+ *β*_*5*_ Race-Black + *β*_*6*_ Race-American Indian or Alaska Native + *β*_*7*_ Race-Asian

+ *β*_*8*_ Race-Hawaiian or Pacific Islander + *β*_*9*_ Race-Other + *β*_*10*_ Race-Two or More Races

+ *β*_*11*_ Ethnicity-Hispanic or Latino + *β*_*12*_ Poverty Ratio + *ε*

#### Model 4

WBI2_Model4_ = *α* + *β*_*1*_ Screen Time/Addiction + *β*_*2*_ Pandemic 2020 + *β*_*3*_ Screen Time/Addiction*Pandemic 2020

+ *β*_*4*_ Elementary School + *β*_*5*_ Junior School + *β*_*6*_ Sex-male + *β*_*7*_ Race-Black

+ *β*_*8*_ Race-American Indian or Alaska Native + *β*_*9*_ Race-Asian

+ *β*_*10*_ Race-Hawaiian or Pacific Islander + *β*_*11*_ Race-Other + *β*_*12*_ Race-Two or More Races

+ *β*_*13*_ Ethnicity-Hispanic or Latino + *β*_*14*_ Poverty Ratio + *ε*

##### Software

SAS version 9.4 (SAS Institute Inc, NC) was used for the analyses described above.

## Results

### Recreational screen time and addiction significantly increased during the pandemic year compared to previous years

The overall demographics of children included in this study show that a large number of elementary (6-7 years old), middle (11-13 years old), and high (14-17 years old) school-age children widely across the US are represented by the NSCH (Table 1). On average, they spent increasing hours of screen recreationally (*i.e*. outside of school work) over the years, with 2.38 hours in 2018, 2.41 hours in 2019, and 2.70 hours in 2020 (p < 0.01 for each of the annual difference) (Table 2). When assessing the proportion of those with likely screen addiction (≥4 hours of recreational screen time a day), we found a significant increase over time, with 22.72% in 2018, 24.39% in 2019, and 32.80% in 2020 (p<0.01 increased compared to each previous year), and especially during the 2020 pandemic year showing a notable 8.41% increase compared to 2019 (Table 2). In general, during the 2020 year, there was a decrease in the percentage of children who spend 2 hours and below on screen compared to previous years in 2018 and 2019, and concomitantly, an increase in the percentage of children who spend 3 hours and above (23.15% in 2018 and 23.79% in 2019, versus 33.86% in 2020), showing an overall changing trend in recreational screen time (p<0.0001) (Table 3).

**Table 1:** Summary of the study population demographics. Categorical demographic variables are shown as n (%) relative to the total population (N=63,211), and the unweighted Poverty Ratio is shown as mean ± standard deviation, with a range of (50 - 400).

|  |  |
| --- | --- |
|  | **N=63,211** |
| **Age group:** |  |
| Elementary school (age 6-10 years) | 21,857(34.57%) |
| Middle school (age 11-13 years) | 15,634 (24.73%) |
| High school (age 14-17 years) | 25,720 (40.69%) |
| **Sex:** |  |
| Male | 32,010 (50.63%) |
| Female | 31,201 (49.37%) |
| **Race:** |  |
| White | 49,209 (77.85%) |
| Black or African American | 4,438 (7.02%) |
| American Indian or Alaska Native | 571 (0.90%) |
| Asian | 3,451 (5.46%) |
| Native Hawaiian or Other Pacific Islander | 352 (0.56%) |
| Some Other Race Alone | 486 (0.77%) |
| Two or More Races | 4,704 (7.44%) |
| **Ethnicity:** |  |
| Hispanic or Latino | 7,452 (11.79%) |
| Otherwise | 55,759 (88.21%) |
| **Poverty Ratio** | 292.44 ± 116.22 |

**Table 2:** Summary of recreational screen time hours, and proportion of screen addiction (≥4 hours) among the different age groups across study years. Student’s t-test p-values are shown for the difference between the value of current versus previous year. Less than one hour of screen time was recorded as 0.5, and four or more was recorded as 4.

|  | Screen time average (hours) |  |  | Percentage of screen addiction<br>(≥4 hours of screen time) |  |  |
| --- | --- | --- | --- | --- | --- | --- |
|  | 2018 | 2019 | 2020 | 2018 | 2019 | 2020 |
| **All ages**<br>**(N=63,211)** | 2.38 | 2.41 | 2.70 | 18,746<br>(22.72%) | 18,212<br>(24.39%) | 26,253<br>(32.80%) |
| *P* value | --- | 0.0045 | <0.0001 | --- | 0.0002 | <0.0001 |
| **Elementary**<br>**school**<br>**(age 6-10 years)**<br>**(n=21,857)** | 1.98 | 2.02 | 2.37 | 6,478<br>(11.78%) | 6,249<br>(13.55%) | 9,130<br>(20.78%) |
| *P* value | --- | 0.0635 | <0.0001 | --- | 0.0026 | <0.0001 |
| **Middle school**<br>**(age 11-13 years)**<br>**(n=15,634)** | 2.46 | 2.47 | 2.79 | 4,561<br>(24.36%) | 4,564<br>(24.30%) | 6,509<br>(34.59%) |
| *P* value | --- | 0.9190 | <0.0001 | --- | 0.9455 | <0.0001 |
| **High school**<br>**(age 14-17 years)**<br>**(n=25,720)** | 2.80 | 2.86 | 3.03 | 7,707<br>(34.88%) | 7,399<br>(38.00%) | 10,614<br>(45.83%) |
| *P* value | --- | 0.0002 | <0.0001 | --- | <0.0001 | <0.0001 |

**Table 3:** Distribution of hours of recreational screen time by year. Chi-square test p-values are shown for the difference between the distribution of the current versus previous year.

|  | < 1 hour | 1 hour | 2 hours | 3 hours | ≥4 hours | *P* value |
| --- | --- | --- | --- | --- | --- | --- |
| **Total**<br>**(N=63,211)** | 3,909<br>(6.18%) | 8,775<br>(13.88%) | 18,863<br>(29.84%) | 14,104<br>(22.31%) | 17,560<br>(27.78%) | --- |
| **2018**<br>**(N=18,746)** | 1,315<br>(7.01%) | 3,144<br>(16.77%) | 5,903<br>(31.49%) | 4,045<br>(21.58%) | 4,339<br>(23.15%) | --- |
| **2019**<br>**(N=18,212)** | 1,372<br>(7.53%) | 2,828<br>(15.53%) | 5,726<br>(31.44%) | 3,954<br>(21.71%) | 4,332<br>(23.79%) | <0.0001 |
| **2020**<br>**(N=26,253)** | 1,222<br>(4.65%) | 2,803<br>(10.68%) | 7,234<br>(27.55%) | 6,105<br>(23.25%) | 8,889<br>(33.86%) | <0.0001 |

### Psychological well-being issue worsened significantly during the pandemic

We developed two well-being issue (WBI) scores as overall measures of children’s psychological mental health conditions. The WBI1 score is a composite score considering answers to six different NCHS survey questions related to the inability to stay calm, arguing behavior, difficulty in making and maintaining friendship, inability to finish tasks, lack of curiosity, and difficulty for parents to care for, and the WBI2 score is a dichotomous score considering whether any of the six conditions, or additionally, depression or anxiety were identified in the survey answers (Supplementary Table S2). While the WBI1 score was not significantly different between 2018 and 2019 (0.843 and 0.839, respectively, p=0.0641), it increased significantly in the pandemic year, 2020 (1.031, p<0.0001 compared to 2019) (Table 4). Similarly, while the WBI2 score was comparable between 2018 and 2019 (47.6% and 45.8%, respectively, p=0.284), it increased significantly in 2020 (51.7%, p<0.0001) (Table 4). These observations demonstrate that the pandemic year had an impact on children’s psychological well-being.

**Table 4:** Changes in the psychological well-being issue scores, WBI1 and WBI2 before (2018 and 2019), and during (2020) the COVID-19 pandemic. The mean and Student’s t-test p-values are shown for WBI1, and the proportions and Chi-square test p-values are shown for the difference between the value of the current versus previous year.

| **WBI1** |  |  |  |
| --- | --- | --- | --- |
|  | **N** | **Mean** | ***P* value** |
| **Total** | 63,211 | 0.905 | --- |
| **2018** | 18,746 | 0.843 | --- |
| **2019** | 18,212 | 0.839 | 0.0641 |
| **2020** | 26,253 | 1.031 | <0.0001 |
| **WBI2** |  |  |  |
|  | **N** | **Proportion (%)** | ***P* value** |
| **Total** | 63,211 | 47.6% | --- |
| **2018** | 18,746 | 45.3% | --- |
| **2019** | 18,212 | 45.8% | 0.284 |
| **2020** | 26,253 | 51.7% | <0.0001 |

### Recreational screen time and screen addiction are associated with psychological well-being issue scores, and the pandemic year significantly contributed to the association

We constructed various multivariate GLS models to understand the association between recreational screen time or addiction (≥4 hours a day) and the WBI1 score, adjusting for age, race, ethnicity, and poverty ratio, first without including information on the pandemic year 2020 (Model 1) and separately, considering the pandemic year 2020 as a covariate in the model or as an interaction term (Model 2) to understand how it may modify the association (Table 5). We found that screen time and screen addiction were both positively associated with WBI1 (p<0.001), in all models. Moreover, in the models that included the covariate for the pandemic year 2020, as well as the interaction term between screen time or addiction and the pandemic year (Model 2), both showed a significant positive coefficient estimate.

**Table 5:**
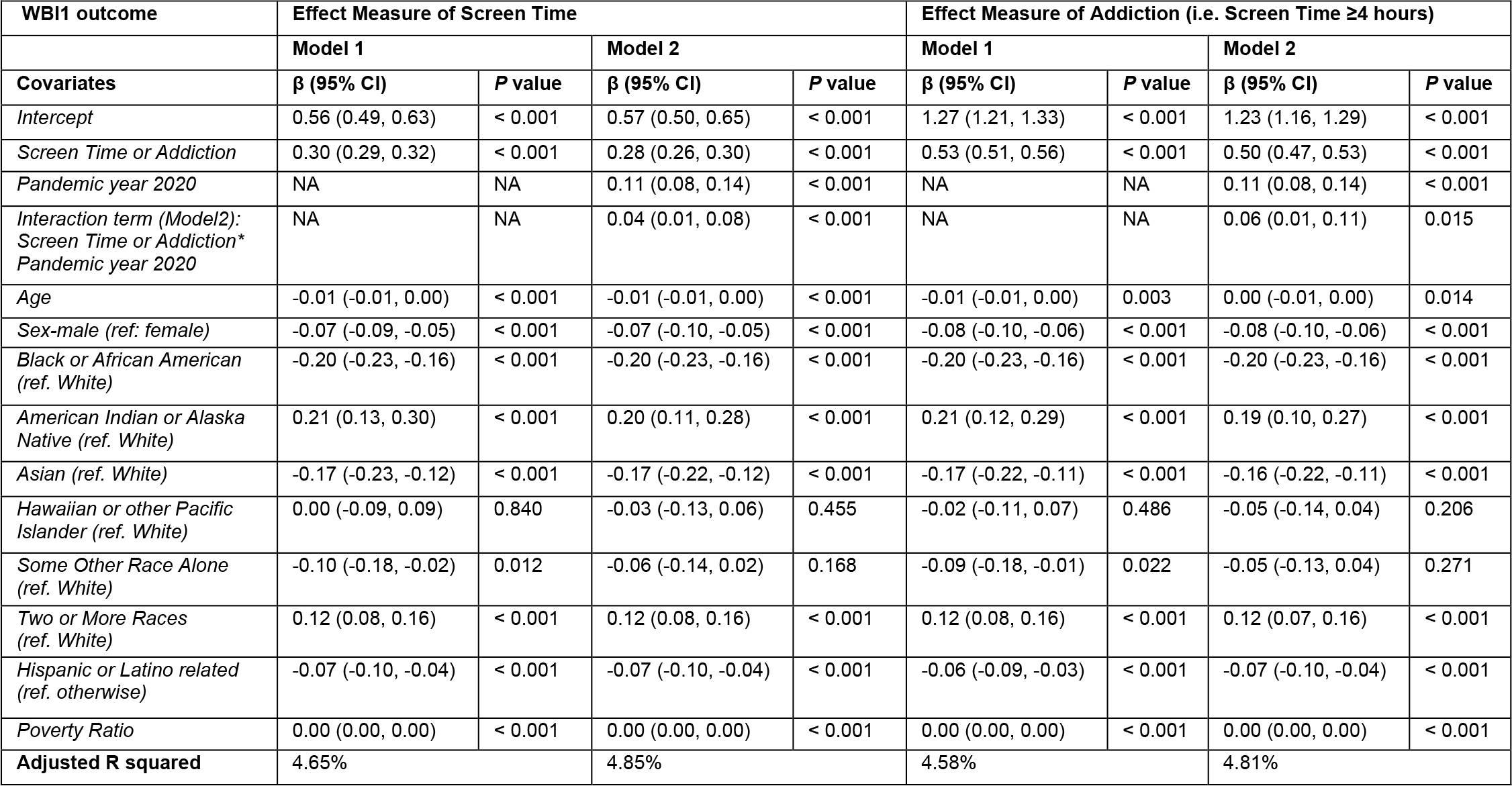
GLS models 1 and 2 regression results for children’s psychological well-being scores WBI1, to assess the impact of hours spent on recreational screen on an average school day (*Screen Time*), and to assess the impact on recreational screen addiction of at least 4 hours on an average school day (*Addiction*). Regression was performed among 50,527 observations of those with at least one hour of screen usage every weekday.

| WBI1 outcome | Effect Measure of Screen Time |  |  |  | Effect Measure of Addiction (i.e. Screen Time ≥4 hours) |  |  |  |
| --- | --- | --- | --- | --- | --- | --- | --- | --- |
|  | Model 1 |  | Model 2 |  | Model 1 |  | Model 2 |  |
| Covariates | β (95% CI) | P value | β (95% CI) | P value | β (95% CI) | P value | β (95% CI) | P value |
| *Intercept* | 0.56 (0.49, 0.63) | < 0.001 | 0.57 (0.50, 0.65) | < 0.001 | 1.27 (1.21, 1.33) | < 0.001 | 1.23 (1.16, 1.29) | < 0.001 |
| *Screen Time or Addiction* | 0.30 (0.29, 0.32) | < 0.001 | 0.28 (0.26, 0.30) | < 0.001 | 0.53 (0.51, 0.56) | < 0.001 | 0.50 (0.47, 0.53) | < 0.001 |
| *Pandemic year 2020* | NA | NA | 0.11 (0.08, 0.14) | < 0.001 | NA | NA | 0.11 (0.08, 0.14) | < 0.001 |
| *Interaction term (Model2):*<br>*Screen Time or Addiction\**<br>*Pandemic year 2020* | NA | NA | 0.04 (0.01, 0.08) | < 0.001 | NA | NA | 0.06 (0.01, 0.11) | 0.015 |
| *Age* | -0.01 (-0.01, 0.00) | < 0.001 | -0.01 (-0.01, 0.00) | < 0.001 | -0.01 (-0.01, 0.00) | 0.003 | 0.00 (-0.01, 0.00) | 0.014 |
| *Sex-male (ref: female)* | -0.07 (-0.09, -0.05) | < 0.001 | -0.07 (-0.10, -0.05) | < 0.001 | -0.08 (-0.10, -0.06) | < 0.001 | -0.08 (-0.10, -0.06) | < 0.001 |
| *Black or African American*<br>*(ref. White)* | -0.20 (-0.23, -0.16) | < 0.001 | -0.20 (-0.23, -0.16) | < 0.001 | -0.20 (-0.23, -0.16) | < 0.001 | -0.20 (-0.23, -0.16) | < 0.001 |
| *American Indian or Alaska*<br>*Native (ref. White)* | 0.21 (0.13, 0.30) | < 0.001 | 0.20 (0.11, 0.28) | < 0.001 | 0.21 (0.12, 0.29) | < 0.001 | 0.19 (0.10, 0.27) | < 0.001 |
| *Asian (ref. White)* | -0.17 (-0.23, -0.12) | < 0.001 | -0.17 (-0.22, -0.12) | < 0.001 | -0.17 (-0.22, -0.11) | < 0.001 | -0.16 (-0.22, -0.11) | < 0.001 |
| *Hawaiian or other Pacific*<br>*Islander (ref. White)* | 0.00 (-0.09, 0.09) | 0.840 | -0.03 (-0.13, 0.06) | 0.455 | -0.02 (-0.11, 0.07) | 0.486 | -0.05 (-0.14, 0.04) | 0.206 |
| *Some Other Race Alone*<br>*(ref. White)* | -0.10 (-0.18, -0.02) | 0.012 | -0.06 (-0.14, 0.02) | 0.168 | -0.09 (-0.18, -0.01) | 0.022 | -0.05 (-0.13, 0.04) | 0.271 |
| *Two or More Races*<br>*(ref. White)* | 0.12 (0.08, 0.16) | < 0.001 | 0.12 (0.08, 0.16) | < 0.001 | 0.12 (0.08, 0.16) | < 0.001 | 0.12 (0.07, 0.16) | < 0.001 |
| *Hispanic or Latino related*<br>*(ref. otherwise)* | -0.07 (-0.10, -0.04) | < 0.001 | -0.07 (-0.10, -0.04) | < 0.001 | -0.06 (-0.09, -0.03) | < 0.001 | -0.07 (-0.10, -0.04) | < 0.001 |
| *Poverty Ratio* | 0.00 (0.00, 0.00) | < 0.001 | 0.00 (0.00, 0.00) | < 0.001 | 0.00 (0.00, 0.00) | < 0.001 | 0.00 (0.00, 0.00) | < 0.001 |
| **Adjusted R squared** | 4.65% |  | 4.85% |  | 4.58% |  | 4.81% |  |

Using a similar approach, we also constructed various multivariate logistic regression models for the outcome of the WBI2 score (Table 6). Adjusting for age group, sex, race, ethnicity, and poverty ratio, each unit increase in screen time was associated with 1.448 times the odds of WBI2 (p<0.001), and screen addiction was associated with 1.980 times the odds of WBI2 (p<0.001), in models without the pandemic year 2020 and interaction terms (Model 3). Including the pandemic year 2020 and its interaction term, the screen hours was associated with 1.448 times the odds, and screen addiction was associated with 1.903 times the odds of WBI2 (Model 4). In these models, the pandemic year 2020 was associated with 1.162 times the odds of WBI2 for the model with screen time, or 1.172 times for the model with addiction. The interaction terms also showed a positive significant association (p<0.001), demonstrating the impact of the pandemic on strengthening the effect of screen time and addiction on reduced psychological well-being.

**Table 6:** The odds ratio (OR) estimates of multivariable logistic regression models for the outcome of children’s psychological well-being score, WBI2, to assess the impact of hours spent on recreational screen on an average school day (*Screen Time*), and to assess the impact of recreational screen addiction of at least 4 hours on an average school day (*Addiction*). Regression was performed among 50,527 observations of those with at least one hour of screen usage every weekday.

| WB12 outcome | Effect Measure of Screen Time |  |  |  | Effect Measure of Addiction (i.e. Screen Time ≥4 hours) |  |  |  |
| --- | --- | --- | --- | --- | --- | --- | --- | --- |
|  | Model 3 |  | Model 4 |  | Model 3 |  | Model 4 |  |
| Covariates | OR (95% CI) | P value | OR (95% CI) | P value | OR (95% CI) | P value | OR (95% CI) | P value |
| *Screen Time or Addiction* | 1.484 (1.483, 1.485) | < 0.001 | 1.448 (1.447, 1.450) | < 0.001 | 1.980 (1.977, 1.983) | < 0.001 | 1.903 (1.899, 1.96) | < 0.001 |
| *Pandemic year 2020* | NA | NA | 1.162 (1.160, 1.164) | < 0.001 | NA | NA | 1.172 (1.170, 1.174) | < 0.001 |
| *Interaction term (Model4):*<br>*Screen Time or Addiction\**<br>*Pandemic year 2020* | NA | NA | 1.049 (1.047, 1.051) | < 0.001 | NA | NA | 1.069 (1.066, 1.072) | < 0.001 |
| *Elementary School*<br>*(ref. High School)* | 1.057 (1.056, 1.059) | < 0.001 | 1.048 (1.046, 1.050) | < 0.001 | 1.037 (1.035, 1.039) | < 0.001 | 1.028 (1.027, 1.030) | < 0.001 |
| *Middle School*<br>*(ref. High School)* | 1.045 (1.043, 1.047) | < 0.001 | 1.039 (1.037, 1.041) | < 0.001 | 1.043 (1.042, 1.045) | < 0.001 | 1.038 (1.036, 1.040) | < 0.001 |
| *Sex-male (ref. female)* | 0.993 (0.991, 0.994) | < 0.001 | 0.994 (0.993, 0.996) | 0.030 | 1.002 (1.001, 1.004) | 0.002 | 1.004 (1.002, 1.005) | < 0.001 |
| *Black or African American*<br>*(ref. White)* | 0.695 (0.694, 0.697) | < 0.001 | 0.696 (0.694, 0.697) | < 0.001 | 0.697 (0.696, 0.699) | < 0.001 | 0.697 (0.696, 0.699) | < 0.001 |
| *American Indian or Alaska*<br>*Native (ref. White)* | 1.152 (1.146, 1.158) | < 0.001 | 1.126 (1.120, 1.132) | < 0.001 | 1.140 (1.134, 1.146) | < 0.001 | 1.113 (1.107, 1.120) | < 0.001 |
| *Asian (ref. White)* | 0.724 (0.722, 0.727) | < 0.001 | 0.725 (0.723, 0.727) | < 0.001 | 0.731 (0.728, 0.733) | < 0.001 | 0.731 (0.729, 0.734) | < 0.001 |
| *Hawaiian or other Pacific*<br>*Islander (ref. White)* | 0.975 (0.970, 0.981) | < 0.001 | 0.938 (0.933, 0.944) | 0.852 | 0.946 (0.941, 0.951) | < 0.001 | 0.908 (0.903, 0.914) | < 0.001 |
| *Some Other Race Alone*<br>*(ref. White)* | 0.934 (0.930, 0.939) | 0.080 | 0.993 (0.988, 0.998) | < 0.001 | 0.944 (0.939, 0.948) | < 0.001 | 1.007 (1.002, 1.012) | < 0.001 |
| *Two or More Races*<br>*(ref. White)* | 1.208 (1.205, 1.211) | < 0.001 | 1.206 (1.203, 1.209) | < 0.001 | 1.206 (1.203, 1.209) | < 0.001 | 1.204 (1.201, 1.207) | < 0.001 |
| *Hispanic or Latino related*<br>*(ref. otherwise)* | 0.904 (0.902, 0.906) | < 0.001 | 0.899 (0.897, 0.900) | < 0.001 | 0.909 (0.908, 0.911) | < 0.001 | 0.903 (0.902, 0.905) | < 0.001 |
| *Poverty Ratio* | 0.999 (0.999, 0.999) | < 0.001 | 0.999 (0.999, 0.999) | < 0.001 | 0.999 (0.999, 0.999) | < 0.001 | 0.999 (0.999, 0.999) | < 0.001 |

Collectively, these various regression results show that higher screen time and addiction are significantly related to low psychological well-being over 2018-2020, and that the pandemic year of 2020 significantly contributed to children’s low psychological well-being. Furthermore, the interaction terms between screen time or addiction and the year 2020 confirms the impact of the pandemic on strengthening the effect of screen time or addiction on children’s psychological well-being.

## Discussion

The NCHS dataset we used for this study included a large number of school-age children across the US, providing results that are applicable widely. Our study found a significant surge in recreational screen time and screen addiction during the 2020 pandemic year, compared to prior years of 2018 and 2019.

Concomitantly, a significant decline in psychological well-being was also detected during 2020, compared to 2018 and 2019. While previous studies explored the impact of the pandemic on the overall (*i.e*. instructional and recreational combined) screen time usage^7,8^, or their associations with mental health among a subset of children with reported severe COVID-related family and economic hardship^9^ or worry^10^ among a narrow age range, they did not include an analysis comparing the years before and during the pandemic, evaluate the effect of recreational screen overuse specifically, or use a large sample of all school-age children widely across the US, as in this study.

We developed various psychological well-being issue (WBI) scores and used them to quantify the association with recreational screen time and addiction by constructing regression models. Furthermore, we also developed additional models to assess the impact of the pandemic itself on psychological well-being, as well as whether it strengthened the effect of screen time and addiction on psychological well-being. Our results demonstrate that screen time and addiction are strongly associated with a decline in psychological well-being, and moreover, that the pandemic was a significant factor independently, and as an effect measure modifier.

Compared to previous studies evaluating the association of screens with children’s mental health during the pandemic only, our study design analyzing years before and during the pandemic allowed additional characterization of the impact of the pandemic as an event alone, highlighting the strength of our results.

Moreover, we evaluated a large number of school-aged children of a wide age range and regardless of economic status, generally across the US, rather than specific groups of children. Additional studies to compare the association after the pandemic, as well as studies in different regions outside of the US could also be informative.

With innovations in online technology and decreasing costs, education and social connections provided through screens have become increasingly more accessible and equitable, providing more opportunities widely.^16^ However, despite the efficient educational information dissemination benefits and entertainment that the internet, computer, device, and television screens offer to the modern world, the number of hours children spend on screens is a major concern.^17^ The COVID-19 pandemic necessitated access to education and social connections through screens, which was vital to the maintenance of interpersonal connections and mental health during pandemic-related lockdowns.^18^ However, as our study’s results suggest, aside from the education and social aspect of screens, it is necessary to be cautious of excessive recreational screen time. Especially among children, hinderance of daily recreational activities involving observations and explorations of their environment by excessive screen time, could negatively impact cognitive development.^19^ In addition to impeding cognitive development, increased screen time by television and social media use in early life are associated with other adverse physiological effects, including weight gain, sleep disruption, depression and inattention problems, and developmental delays. ^20,21^ It has been previously suggested that while excessive internet use is well-established to be associated with depression, too little usage hours could also be related to depression^20^, which is also suggested from our study that found that screen less that one hour of screen time resulted in worse well-being compared to one hour. Taken together, it may suggest that optimal screen time exposure that allows sufficient remote social interactions and entertainment, while limiting excess use is important, and further studies to identify the optimal length of time that is advantageous for psychological well-being is informative.

This study has several limitations pertaining to the use of the NSCH dataset used and analysis methods. The 2020 data were collected between June 2020 and January 2021^22^, during which most children were quarantined and undergoing online instruction, however, it is not possible to determine the exact time frame when the study participants responded to the survey questions and there may be variability in the phase of the pandemic. Moreover, the survey answer choices on recreational screen hours are designed such that four or more hours is the maximum recorded response, and anything above is classified as the same answer. The response is also according to self-reported hours on average on a weekday, although daily hours is expected to be highly variable, in reality. Furthermore, the NSCH questionnaire variables related to psychological well-being used in this study include some that were reported by the parents. Another limitation is that currently, there is no established definition for screen addiction. Addiction is characterized by psychological dependency and overuse^23,24,25,26,27^, and accordingly in our study, we considered reported use of at least four hours of recreational (*i.e*. non-school related) screen time during school days as excessive use, as it suggests that a child is spending all the time left in a day outside of sleep, school work, commute and feeding time, on a screen. It is not possible to ascertain whether these children have psychological dependency on using screen devices.^28^ Future studies with survey questions directly related to screen addiction would be beneficial. Finally, compared to surveys administered before the pandemic when children were physically in school, self-reported recreational screen time outside of school-related activities during the pandemic when online instruction was ongoing, may be more likely to yield overestimation errors.

Despite the limitations above, the results of this study showing strong associations between screen time and psychological well-being issues, as well as the significant impact of the pandemic in this association, using a large survey dataset collected widely across the US is informative. We demonstrate annual increases in U.S. school-age children’s recreational screen time and psychological well-being between 2018 and 2020, and provide evidence of the 2020 pandemic as an independent factor in worsening US school-age children’s screen time and well-being conditions, as well as strengthens the association. Our results may suggest that prevention of screen overuse and addiction among school-age children by incorporating other activities such as physical activity, sports, music, arts, and social hours^7,28,29,30^ off screens may improve psychological well-being.

## Data Availability

All data used in this study are publicly available and were accessed from the National Survey of Children's Health (NSCH) website (https://www.childhealthdata.org/learn-about-the-nsch/NSCH)

## Abbreviations

NSCH: National Survey of Children’s Health
WB: well-being
WBI: well-being issue
COVID-19: coronavirus disease of 2019

## Author Contributions

HTW originally conceived the study and collected the datasets. HTW and JL performed the formal analyses. HTW drafted the first draft. HTW, JL, and AT interpreted the data, edited the manuscript, and approved the final version.

## Data statement

All data used in the manuscript are publicly available datasets.

