## Supplementary Materials for "The Change of Screen Time and Screen Addiction, and their Association with Psychological Well-being During the COVID-19 Pandemic: An Analysis of US Country-Wide School-Age Children and Adolescents Between 2018 and 2020"

| **NSCH survey variable (variable name according to the public use data file) and detailed description of the question and answer choices on the original survey** | **Recoded variable used in this study** |
|---|---|
| **Recreational screen time (SCREENTIME).**<br>“On most week days, about how much time did this child spend in front of a TV, computer, cellphone or other electronic device watching programs, playing games, accessing the internet or using social media, not including school work?”<br>Answer choices: 1 = Less than 1 hour, 2 = 1 hour, 3 = 2 hours, 4 = 3 hours or 5 = 4 or more hours. | ***Screen Time***<br>0.5 = SCREENTIME of 1 (*i.e.* Less than 1 hour),<br>1 = SCREENTIME of 2 (*i.e.* 1 hour),<br>2 = SCREENTIME of 3 (*i.e.* 2 hours),<br>3 = SCREENTIME of 4 (*i.e.* 3 hours),<br>4 = SCREENTIME of 5 (*i.e.* 4 or more hours). |
| **Ability to remain calm and in control when challenged (K7Q85_R).**<br>“How often: Does this child stay calm and in control when faced with a challenge?”<br>Answer choices: 1 = Always, 2 = Usually, 3 = Sometimes, 4 = Never. | ***Not Calm***<br>1 = K7Q85_R value of 3 or 4, indicating the child does not stay calm or in control,<br>0 = K7Q85_R value of 1 or 2, indicating the child does stay calm or in control. |
| **Children who argue too much (K7Q70_R).**<br>“How often: Does this child argue too much?”<br>Answer choices: 1 = Always, 2 = Usually, 3 = Sometimes, 4 = Never. | ***Argues Too Much***<br>1 = K7Q70_R1 value of 1 or 2, indicating the child argues too much,<br>0 = K7Q70_R1 value of 3 or 4, indicating the child does not argue too much. |
| **Difficulty making or keeping friends (MAKEFRIEND).**<br>“Compared to other children his or her age, how much difficulty does this child have making or keeping friends?”<br>Answer choices: 3 = A lot of difficulty, 2 = A little difficulty, 1 = No difficulty. | ***Difficult to Make Friends***<br>1 = MAKEFRIEND of 2 or 3, indicating the child has difficulty making or keeping friends,<br>0 = MAKEFRIEND of 1, indicating the child does not have difficulty making or keeping friends. |
| **Children who work to finish the tasks they start (K7Q84_R).**<br>“How often: Does this child work to finish tasks they start?”<br>Answer choices: 1 = Always, 2 = Usually, 3 = Sometimes, 4 = Never. | ***Does Not Finish Tasks***<br>1 = K7Q84_R value of 3 or 4, indicating the child does not finish tasks,<br>0 = K7Q84_R value of 1 or 2, indicating the child does finish tasks. |
| **Children who show interest and curiosity in learning new things (K6Q71_R).**<br>“How often: Does this child show interest and curiosity in learning new things?”<br>Answers choices: 1 = Always, 2 = Usually, 3 = Sometimes, 4 = Never. | ***Not Curious***<br>1 = K6Q71_R value of 3 or 4, indicating the child does not show interest or curiosity,<br>0 = K6Q71_R value of 1 or 2, indicating the child does show interest and curiosity. |
| **Parent felt a child is difficulty to care for (K8Q31).**<br>“During the past month, how often have you felt: that this child is much harder to care for than most children his or her age?”<br>Answers choices: 1 = Never, 2 = Rarely, 3 = Sometimes, 4 = Usually, 5 = Always. | ***Difficult to Care***<br>1 = K8Q31 value of 3, 4 or 5, indicating the child is difficult to care for,<br>0 = K8Q31 value of 1 or 2, indicating the child is not difficult to care for. |
| **Been diagnosed with depression (K2Q32A)**<br>“Has a doctor or other health care provider EVER told you that this child has depression?” Answers choices: 1 = Yes, 2 = No. | ***Depression***<br>1 = K2Q32A value of 1, indicating a doctor or other health care provider has diagnosed that the child has depression,<br>0 = K2Q32A value of 2, indicating the child has not been diagnosed with depression. |
| **Been diagnosed with anxiety (K2Q33A)**<br>“Has a doctor or other health care provider EVER told you that this child has anxiety?” Answers choices:1 = Yes, 2 = No. | ***Anxiety***<br>1 = K2Q33A value of 1, indicating the child ever had anxiety,<br>0 = K2Q33A value of 2, indicating the child has never had anxiety. |

**Table S1: Description of the survey questions and answer choices from the NSCH, and recoded variables used for the analyses in our study.**

| **Well-being Issue (WBI) Score** | **Values** | **Definition** |
|---|---|---|
| WBI1 | 0 - 6 | Sum of six WBI symptom dichotomous variables excluding depression and anxiety (*Not Calm, Argues Too Much*, *Difficult to Make Friends*, *Does Not Finish Tasks, Not Curious*, and *Difficult to Care*) |
| WBI2 | 1 or 0 | 1 = has at least one of the eight WBI symptoms (*Not Calm*, *Argues Too Much*, *Difficult to Make Friends*, *Does Not Finish Tasks, Not Curious*, *Difficult to Care*, *Depression*, or *Anxiety*), or<br>0=none of the symptoms |

**Table S2: Description of the construction of the well-being scores, WBI1 and WBI2.**

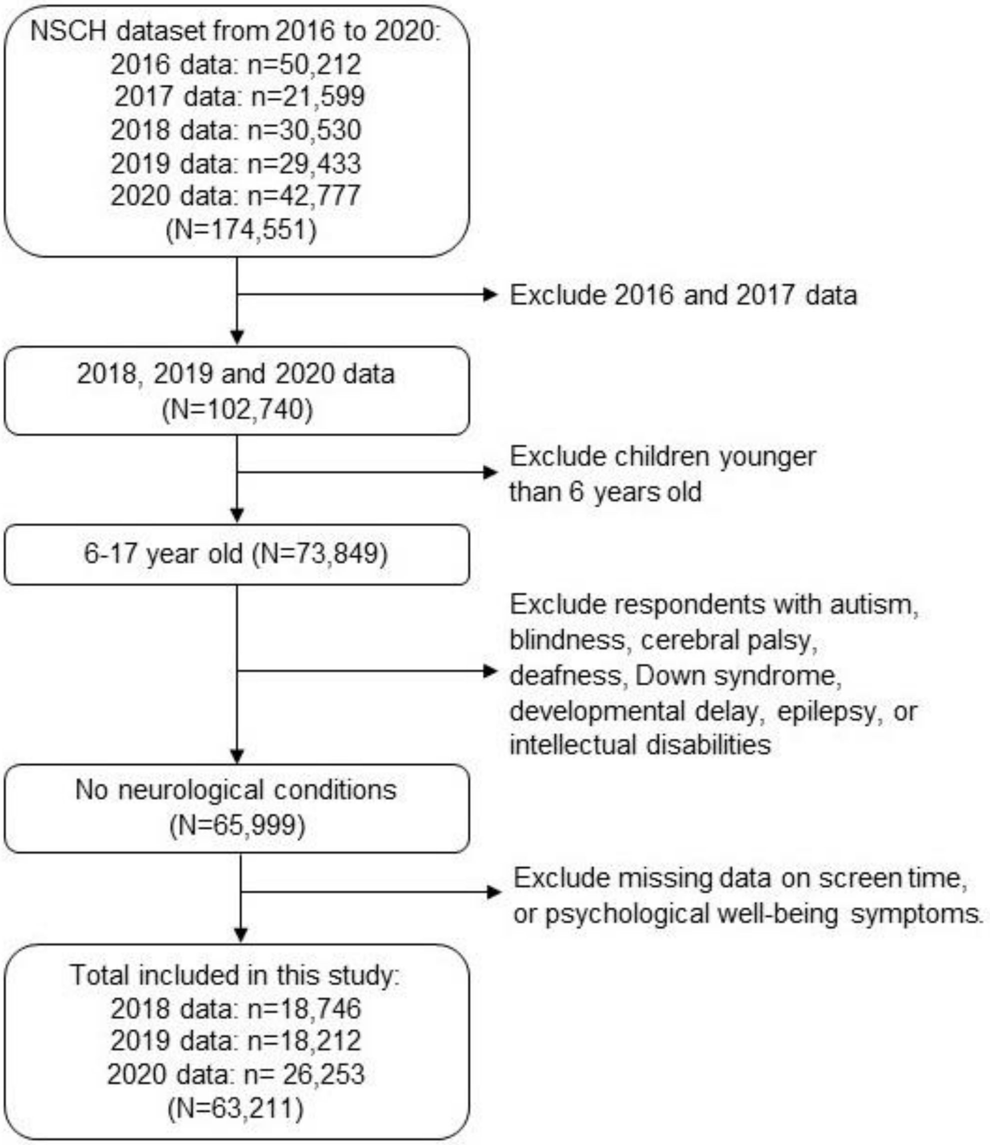


**Figure S1: Flowchart of study inclusion criteria.**

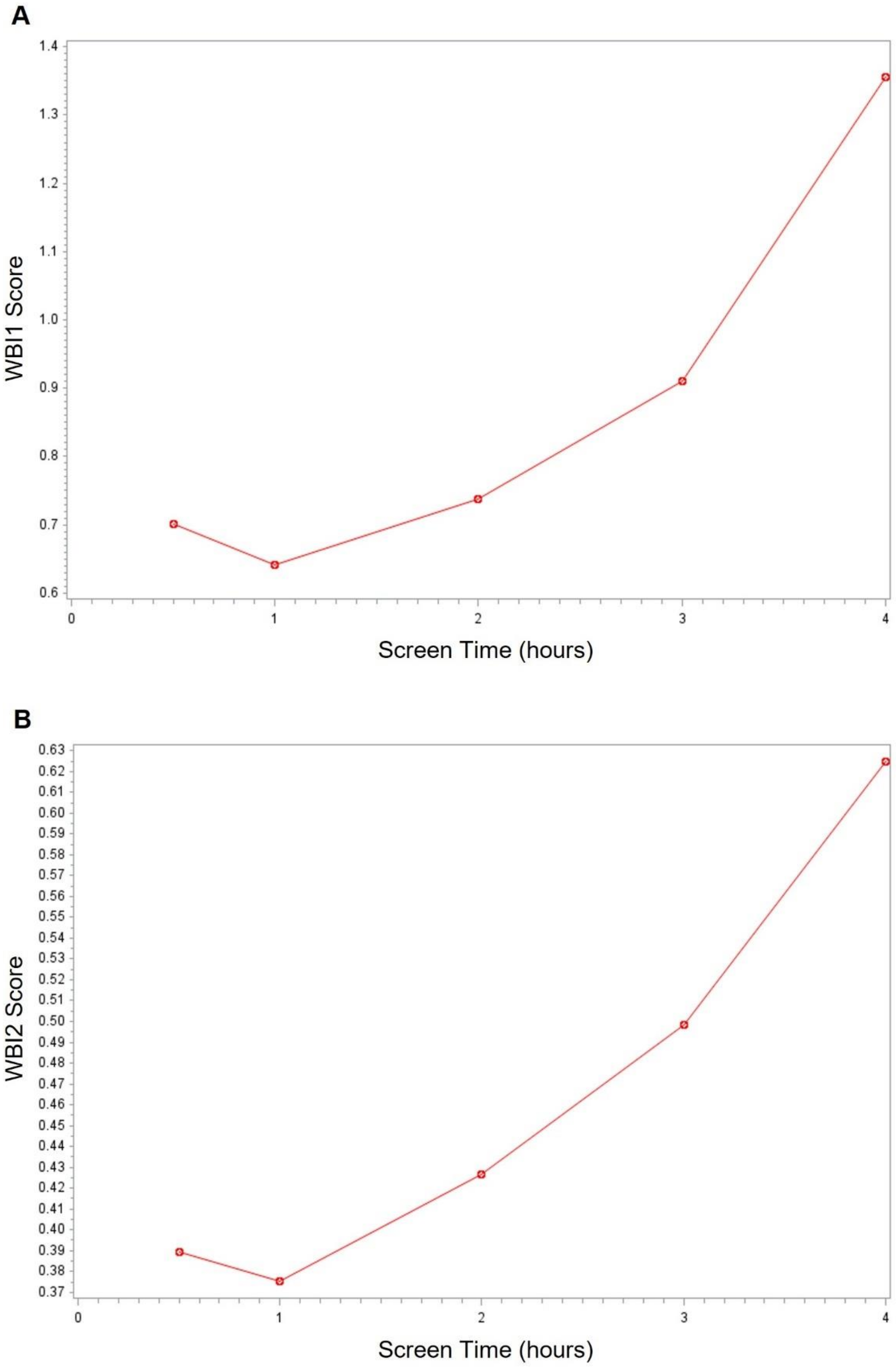


**Figure S2: Assessment of recreational screen time (x-axis) by the psychological average well-being scores (y-axis) for (A) WBI1 and (B) WBI2**
